# Integrating Clustering and Semantic Similarity for MAUDE Database Dimensionality Reduction

**DOI:** 10.1101/2024.12.03.24318439

**Authors:** Lei Hua

## Abstract

**Objective:** To develop and evaluate an automated methodology for dimensionality reduction of the FDA’s MAUDE database through schema matching and merging.

**Methods:** We conducted 96 trails integrating clustering algorithms with semantic similarity evaluations using the DeepSeek V2.5 API. This approach identified and merged semantically similar tables. Feature extraction was performed using Term Frequency-Inverse Document Frequency (TF-IDF) vectorization and Sentence Transformer embeddings. The methodology was assessed against manual groupings provided by domain experts using metrics such as Adjusted Rand Index (ARI), Normalized Mutual Information (NMI), precision, recall, and F1 score. Different similarity thresholds (0.7, 0.8, 0.9) were applied to evaluate their impact on table merging performance.

**Results:** The integration of clustering with semantic similarity evaluations enhanced the F1 score from 0.51 (clustering alone) to 1.00, utilizing fewer than 1,425 API similarity evaluations. Consequently, the number of tables was compressed from 113 to 13–16 table groups, a reduction of 86% to 89%. In addition, the application of clustering algorithms decreased the number of table pair comparisons by 77% to 83%. Sentence Transformer embeddings outperformed TF-IDF vectorization in clustering performance, with F1 scores increasing from a range of approximately 0.51–0.87 to 0.51–0.95 in clustering-only scenarios. DeepSeek V2.5 demonstrated the potential to match and quantify subtle semantic differences across various similarity thresholds, maintaining high merging accuracy with F1 scores reaching up to 1.00.

**Conclusion:** The proposed automated dimensionality reduction methodology effectively enhances data quality and analysis efficiency within the MAUDE database. By reducing the number of tables to manageable groups, optimizing context lengths, and leveraging DeepSeek V2.5’s semantic matching capabilities, the framework streamlines data processing and ensures compatibility with advanced analytical tools such as Large Language Models (LLMs). This makes the methodology applicable across various industries, facilitating more efficient and accurate data analysis workflows

## 1. Introduction

Analyzing the FDA’s Manufacturer and User Facility Device Experience (MAUDE) database is crucial for post-market surveillance of medical device safety and effectiveness, informing regulatory decisions and enhancing patient safety^1^. The MAUDE database is complex. It comprises numerous tables with inconsistent schemas and non-standardized naming conventions, presenting substantial challenges for researchers, particularly those new to the field or with limited computational expertise^2^. High personnel turnover and inadequate documentation of systems further complicate data understanding and processing^3,4^. Without a thorough understanding of data structures and semantic relationships, effective data analysis and extraction become nearly impossible, leading to inefficiencies and potential inaccuracies in research outcomes^5^.

These challenges are not unique to the MAUDE database but are common in historical data analysis scenarios, such as integrating institutional legacy data for secondary use. Even experienced Extract, Transform, Load (ETL) professionals invest significant time deciphering intricate data structures^6^. They must understand implicit relationships among databases, tables, and fields to ensure data integrity. Effective ETL processes require deep data comprehension to avoid redundancy, minimize rework, and ensure accurate data transformation. In this context, schema matching and de-redundancy are crucial strategies. They reduce the complexity of data integration tasks and streamline the overall data processing workflow^7,8^. These methods help align disparate data sources by appropriately grouping similar tables and eliminating unnecessary duplicates, enabling more efficient and reliable analysis.

Recent advancements in Large Language Models (LLMs) have significantly improved data analysis capabilities, offering enhanced data understanding and interpretation^9,10^. However, LLMs have limitations due to context window sizes; mainstream model APIs support up to 128k tokens, and application dialogue windows accommodate up to 32k tokens. These limitations necessitate careful matching and compression of table structures and example data to maintain context during data chunking. Thus, context compression is critical to preserving semantic relationships, enabling accurate and meaningful analysis outcomes^11^.

In this study, we present a novel framework for dimensionality reduction designed to efficiently navigate and compress the schemas of the MAUDE database. Our approach combines clustering algorithms with semantic similarity evaluations to automate schema matching and eliminate redundancies. Specifically, we employ clustering methods such as K-Means ^12^, hierarchical clustering ^13^, and DBSCAN ^14^ to initially group schemas based on their structural features. Subsequently, DeepSeek V2.5^15^, a pretrained large language model, is utilized to further subdivide these groups through semantic relationship assessments.

The framework enhances the precision of schema grouping while reducing computational overhead. Through the reduction of redundancy and the preservation of essential contextual information, our framework empowers researchers and data scientists to conduct analyses with improved accuracy and efficiency. This leads to the rapid identification of potential issues with medical devices, thereby enhancing patient safety and supporting more timely regulatory decisions.

## 2. Background

Database schema matching and grouping have long been fundamental processes in data integration, data warehousing, and data cleaning. These processes are essential when handling heterogeneous and distributed data sources, where inconsistencies and discrepancies in schemas and formats are prevalent. Over the past decades, numerous frameworks and techniques have been developed to address the challenges associated with schema matching, entity resolution, and data consolidation.

### 2.1 Schema Matching and Mapping

Schema matching is the cornerstone of integrating diverse data sources, involving the identification of correspondences between schema elements of different databases.^16^ Approaches to schema matching are primarily divided into element-level and structure-level matching. Element-level matching focuses on comparing individual schema elements based on attributes such as names, data types, constraints, and values, while structure-level matching examines the relationships between schema elements, including hierarchies and foreign keys. To improve matching accuracy, techniques leverage linguistic similarity, structural similarity, and constraint-based matching. Hybrid approaches that integrate multiple techniques, including machine learning algorithms, have proven particularly effective.^7,8,17^ The methods use schema information and data instances to derive matching rules, thereby automating and refining the schema matching process. Machine learning-based schema matching employs feature selection to train classifiers on attributes like names, data types, and value patterns, significantly enhancing the efficiency and accuracy of schema alignment.

### 2.5 Schema Merging and Data Fusion

Building upon schema matching, schema merging and data fusion involve integrating matched schemas from multiple sources to create a unified global schema.^18,19^ Schema merging frameworks aim to consolidate the structures of source schemas, addressing challenges such as naming conflicts, data type mismatches, and structural differences. Various merging strategies, including intersection, union, and integration of schemas, are employed to achieve a coherent global schema. Effective schema conflict resolution is critical, requiring methodologies that can handle discrepancies and ensure consistency across the integrated schema. Automated merging systems like Cupid ^19^ combine multiple matching techniques, including linguistic and structural methods, and utilize statistical analysis to accurately align schema elements from diverse sources.

### 2.2 Entity Resolution and Record Linkage

Entity resolution, also known as record linkage or deduplication, involves identifying and merging records that refer to the same real-world entity across different data sources. Elmagarmid et al.^20^ surveyed various techniques for duplicate record detection, highlighting the critical role of similarity functions, indexing methods, and machine learning algorithms in this domain. Similarity functions, such as edit distance, Jaccard similarity, and term frequency-inverse document frequency (TF-IDF), are employed to compare textual data and quantify the likeness between records. Indexing methods like blocking and canopy clustering are utilized to reduce the computational complexity by limiting the number of record comparisons. Machine learning algorithms, both supervised and unsupervised, are applied to classify record pairs as matches or non-matches, thereby improving the efficiency and accuracy of entity resolution processes, which are vital for data consolidation.

### 2.4 Clustering and Semantic Similarity for Enhanced Data Integration

Clustering algorithms play a fundamental role in data integration by grouping similar schema elements or tables, thereby facilitating the matching and merging processes.^21,22^ By organizing schema elements into meaningful clusters based on similarity measures prior to matching, clustering significantly reduces the search space for potential matches. This reduction enhances scalability and efficiency, particularly when dealing with large and complex schemas. Tools such as COMA++ ^22^ demonstrate the effectiveness of clustering techniques in optimizing the schema matching process by systematically organizing schema elements into coherent groups, thereby managing extensive and intricate schemas more effectively.

Building upon clustering, semantic similarity measures further augment data integration by capturing the contextual meanings of data elements.^23,24^ Techniques that leverage natural language processing (NLP) and ontologies are essential for comprehending the semantics of schema labels and data values. Approaches that integrate lexical and structural similarity, utilizing resources such as WordNet^25^, enable the computation of semantic distances between schema elements. This semantic enrichment improves matching results by considering the meanings and relationships of terms beyond their syntactic representations. The advancement of deep learning has further refined these semantic similarity computations through the use of word embeddings and language models, which represent words and phrases in continuous vector spaces that encode rich semantic information. These deep learning-based techniques capture contextual nuances, thereby enhancing the accuracy of schema matching and entity resolution by addressing intricate semantic relationships.

The integration of clustering and semantic similarity techniques provides a collaborative approach to data integration. Clustering reduces computational complexity by limiting potential matches, while semantic similarity ensures that these matches are contextually and functionally coherent. This combination leverages clustering’s scalability for large datasets and semantic similarity’s precision through deeper contextual understanding. The unified use of these techniques enhances the efficiency and accuracy of schema matching and entity resolution, facilitating more effective data consolidation and interoperability.

## 3. Materials and Methods

To navigate and analyze the MAUDE database, we developed the Semantic Clustering and Merging Framework (SCMF) for dimensionality reduction, as illustrated in Figure 1. The framework begins with data extraction, retrieving relevant table information from the MAUDE database. Following extraction, feature extraction generates descriptive metadata and identifies key characteristics of each table. These features are then utilized to apply clustering techniques, grouping similar tables into initial clusters based on their attributes. To refine these clusters, we evaluate table similarities using both structural and semantic measures, enabling the merging of highly similar tables and reducing redundancy within the database architecture. Finally, we assess the framework’s effectiveness by comparing the automated groupings with classifications provided by domain experts. This integrated approach of clustering and similarity assessment enhances data processing efficiency and deepens the understanding of underlying data relationships. All SCMF codes, experimental data, intermediate processes, and detailed documentation have been open-sourced under the MIT license and are accessible at https://github.com/leiMizzou/Maude-Schema-Analysis.

**Figure 1.**
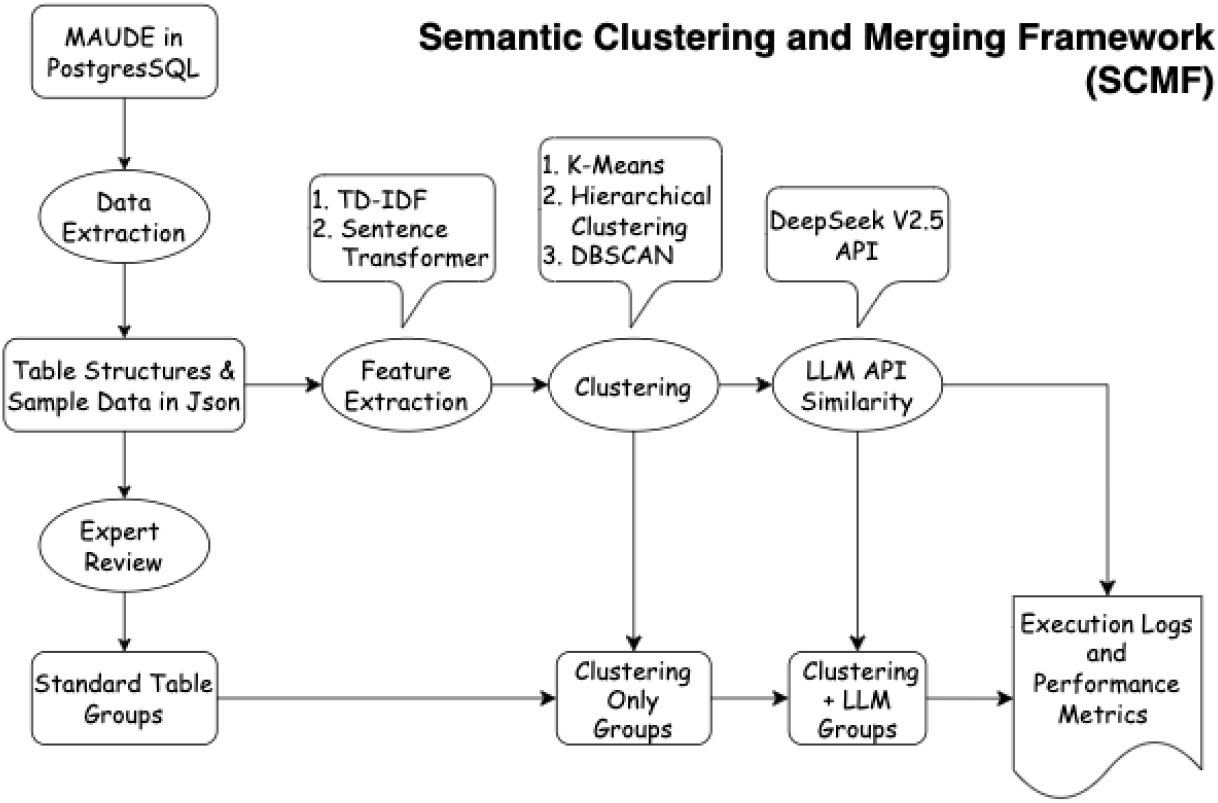
The Workflow of Schema Grouping and Performance Evaluation

### 3.1 System Configuration

The computational environment was optimized for efficient data processing and dimensionality reduction. We utilized Ubuntu 20.04 LTS as the operating system and managed the MAUDE data with PostgreSQL 12.18, ensuring robust handling of large-scale relational data and efficient querying. For computationally intensive tasks such as feature extraction and clustering, Nvidia RTX 3090 GPUs with 24 GB of memory were employed. Semantic similarity computations were performed using the DeepSeek V2.5 API, an open-source, high-performance generative pretrained model.^15^ All experimental code, data analysis, and figure generation were implemented in Python 3.9, leveraging its comprehensive libraries for data science and machine learning.

### 3.2 Dataset

The dataset for this study was sourced from the U.S. Food and Drug Administration’s Manufacturer and User Facility Device Experience (MAUDE) database^i^, encompassing over 40 million records from 1991 to October 2023. These records are distributed across 113 tables, which include 111 MAUDE tables available for download, and 2 self-built reference tables derived from database analysis. The inclusion of self-built tables introduces additional complexity to the analysis process.

To evaluate the performance of dimensionality reduction, these 113 tables were analyzed and grouped by domain experts into semantic groups. This expert-driven grouping serves as the reference standard against which the automated merging results are compared. The table names and their corresponding groupings are summarized in Appendix A.

### 3.3 Data and Feature Extraction

Data extraction involved downloading all compressed MAUDE database files from designated web pages, decompressing them into .txt and .csv formats, and importing them into PostgreSQL 12.18. To ensure data integrity, the row and column counts of each imported table were verified against MAUDE specifications. Each table was then converted to a JSON file to encapsulate its structure and sample data, facilitating subsequent feature extraction and similarity analysis. Feature extraction aimed to capture both structural and semantic characteristics to enhance clustering performance. We employed TF-IDF vectorization^26^ and Sentence Transformer embeddings all-MiniLM-L6-v2^27^ for semantic features. Additionally, numerical features such as the number of columns, counts of primary and foreign keys, and data type distributions were extracted. By experimenting with various feature combinations, we identified the most effective sets for improving clustering accuracy.

### 3.4 Table Clustering and Similarity Computation

Table clustering was undertaken to group similar MAUDE database tables based on their extracted features, revealing inherent relationships and patterns within the dataset. Utilizing a comprehensive feature set that includes both textual (TF-IDF vectorization and Sentence Transformer embeddings) and structural attributes (e.g., number of columns, primary and foreign keys), we applied clustering algorithms such as K-Means^12^ (with both manually specified and automatically determined cluster counts), Hierarchical Clustering^13^, and DBSCAN^14^ to categorize the tables effectively. Given that clustering typically incurs lower computational costs compared to large-scale model inference, initial groupings were rapidly achieved through combinations of TF-IDF and Sentence Transformer features with these algorithms.

To refine the clusters and identify tables suitable for merging, similarity computations were conducted. Initially, Jaccard similarity coefficients^28^ were calculated based on sets of field names to quantify structural overlap, applying a threshold of 0.1 to exclude minimally similar table pairs. Subsequently, the DeepSeek V2.5 API was employed to compute semantic similarity scores between table descriptions, facilitating the detection of subtle semantic differences. Based on these scores, thresholds of 0.7, 0.8, and 0.9 were used to determine whether tables should be merged. The merge strategy prioritized retaining the data and cases from the table with the highest data volume as the representative, effectively consolidating redundant or highly similar tables while preserving the most comprehensive dataset.

The effectiveness of the clustering and similarity computations was evaluated by comparing the automated groupings with manual classifications provided by domain experts. Evaluation metrics including Adjusted Rand Index (ARI)^29^, Normalized Mutual Information (NMI)^30^, Precision, Recall, and F1-Score were employed to quantify the alignment between the automated and manual clusters, ensuring the reliability and accuracy of the SCMF.

Adjusted Rand Index (ARI) measures the similarity between two clustering results by considering all pairs of samples and counting pairs that are assigned in the same or different clusters in the predicted and true clustering, while adjusting for chance groupings. The formula for ARI is:

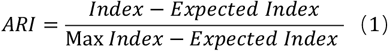

Where *Index* represents the number of agreements between two clustering, *Expected Index* is the expected number of agreements due to random chance, and Max *Index* is the maximum possible value of the Index. ARI ranges from -1 to 1, where a value of 1 indicates perfect agreement between the clustering results, 0 indicates random agreement, and negative values indicate less agreement than expected by chance.

Normalized Mutual Information (NMI) quantifies the mutual dependence between the two clustering, effectively measuring the amount of shared information. The formula for NMI is

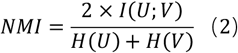

Where *I*(*U*; *V*) is the mutual information between the two clustering *U* and *V*, quantifying the amount of shared information between them. The terms *H*(*U*) and *H*(*V*) denote the entropies of the clustering *U* and *V*, measuring the uncertainty or randomness within each clustering. By incorporating both mutual information and entropy, NMI metric adjusts for the complexity of the clustering, providing a balanced measure of similarity. Its values range from 0 to 1, with higher values indicating a greater degree of similarity between the clustering results.

Precision, Recall, and F1-Score are metrics derived from the classification of sample pairs as either belonging to the same cluster or not. Precision measures the proportion of correctly identified positive pairs (pairs correctly clustered together) out of all pairs identified as positive by the model.

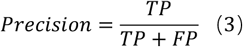

Recall measures the proportion of correctly identified positive pairs out of all actual positive pairs.

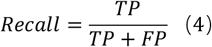

F1-Score is the harmonic mean of Precision and Recall, providing a balance between the two.

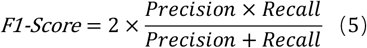

In this context, True Positives (TP) refer to the number of table pairs correctly clustered together in both the automated and manual groupings; False Positives (FP) are the table pairs clustered together by the automated method but not by the manual grouping; and False Negatives (FN) are the table pairs clustered together by the manual grouping but not by the automated method.

### 3.5 Token Efficiency Assessment

To evaluate the effectiveness of the Semantic Clustering and Merging Framework (SCMF) in optimizing the MAUDE database structure, we conducted a token efficiency assessment. This assessment measured the reduction in token usage resulting from the merging of similar tables, which is essential for enhancing data processing efficiency and reducing computational costs.

The assessment began by integrating contextual information from the context.txt file obtained from the FDA MAUDE website.^1^ This file provided comprehensive background information necessary for generating meaningful prompts that encapsulate both the database documentation and its structural details. Two prompt files were created: prompt_before_merging.txt and prompt_after_merging.txt. The former combined context.txt with descriptions of all 113 pre-merged MAUDE database tables, establishing a baseline for token counts before merging. The latter integrated context.txt with descriptions of the post-merged tables produced by the SCMF.

Utilizing the tiktoken^31^ library with the gpt2^32^ encoding scheme, we counted the tokens in each prompt file. To isolate the token counts attributable to the table descriptions, the token count of context.txt was subtracted from each prompt’s total. This step ensured that the token reduction was solely due to the merging process, unaffected by the constant contextual information.

The reduction analysis involved determining both the absolute and percentage decrease in tokens from table descriptions before and after merging. By comparing these metrics, we quantified the extent to which the SCMF streamlined the database schema, thereby enhancing token efficiency.

## 4. Results

The comprehensive analysis, conducted through 96 experimental trails, successfully reduced the number of tables from 113 to approximately 14–16 groups. This reduction highlights the methodology’s effectiveness in dimensionality reduction, facilitating efficient manual analysis and ensuring compatibility with input constraints for Large Language Models (LLMs) and API chat prompts.

### 4.1 SCMF Performance Evaluation

The performance evaluation revealed that applying clustering alone resulted in Adjusted Rand Index (ARI) values ranging from 0.31 to 0.93, Normalized Mutual Information (NMI) values from 0.50 to 0.93, and F1 Scores from 0.51 to 0.95 (Table 4.1, Figure 2A). When clustering was combined with API matching using the DeepSeek V2.5 API, ARI values improved to between 0.85 and 1.00, NMI values to between 0.93 and 1.00, and F1 Scores to between 0.88 and 1.00 (Table 4.1, Figure 2B).

**Table 1.** Comprehensive Performance Metrics and Feature Extraction.

| Category | Method | ARI | NMI | F1 Score | Figure |
| --- | --- | --- | --- | --- | --- |
| **Clustering Only** | Overall Performance | 0.31 – 0.93 | 0.50 – 0.93 | 0.51 – 0.95 | 2A |
| **Clustering + API** | Overall Performance | 0.85 – 1.00 | 0.93 – 1.00 | 0.88 – 1.00 | 2B |
| **Clustering Only**<br>**Feature Extraction** | TF-IDF Vectorization | 0.31 – 0.84 | 0.50 – 0.88 | 0.51 – 0.87 | 2C |
| **Clustering Only**<br>**Feature Extraction** | Transformer Embeddings | 0.31 – 0.93 | 0.50 – 0.93 | 0.51 – 0.95 | 2C |
| **Clustering + API**<br>**Feature Extraction** | TF-IDF Vectorization | 0.94 – 1.00 | 0.97 – 1.00 | 0.95 – 1.00 | 2D |
| **Clustering + API**<br>**Feature Extraction** | Transformer Embeddings | 0.85 – 1.00 | 0.93 – 1.00 | 0.88 – 1.00 | 2D |

**Figure 2.**
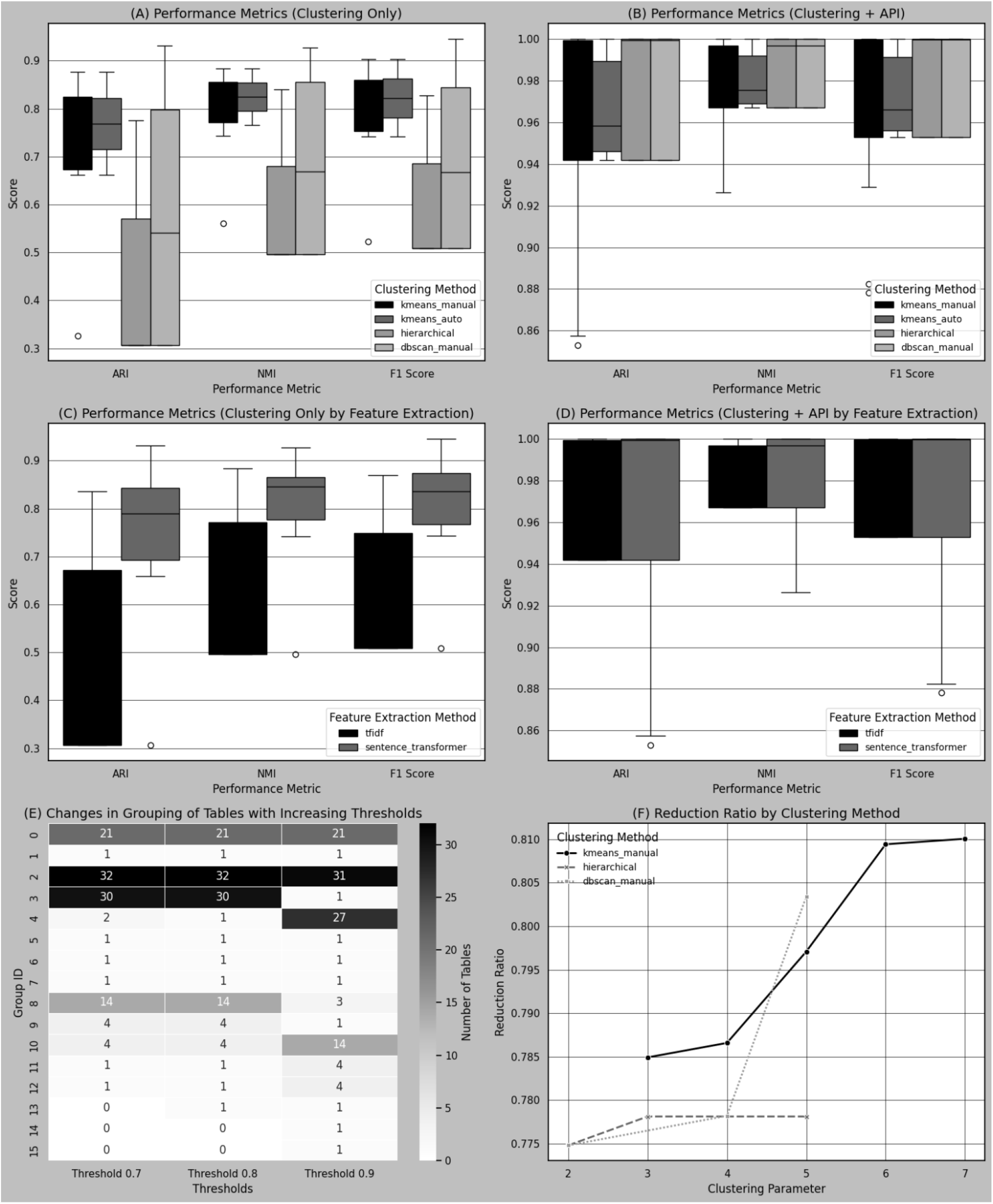
Compilation of Study Results: Performance Metrics, Grouping Refinements, and Reduction ratio

Regarding feature extraction methods, TF-IDF Vectorization achieved ARI values from 0.31 to 0.84, NMI values from 0.50 to 0.88, and F1 Scores from 0.51 to 0.87, whereas Sentence Transformer Embeddings achieved ARI values from 0.31 to 0.93, NMI values from 0.50 to 0.93, and F1 Scores from 0.51 to 0.95 (Table 4.1, Figure 2C). When combined with API matching, TF-IDF Vectorization yielded ARI values from 0.94 to 1.00, NMI values from 0.97 to 1.00, and F1 Scores from 0.95 to 1.00, while Sentence Transformer Embeddings achieved ARI values from 0.85 to 1.00, NMI values from 0.93 to 1.00, and F1 Scores from 0.88 to 1.00 (Table 4.1, Figure 2D).

**Table 2.** Similarity Threshold Effects.

| Similarity Threshold | Effect on Groupings | Num of Groups | Figure |
| --- | --- | --- | --- |
| 0.7 | More merged groups | 13 | 2E |
| 0.8 | Balanced merging and refinement of groups | 14 | 2E |
| 0.9 | Stricter similarity requirements, more refined groupings | 16 | 2E |

The analysis of similarity thresholds showed that increasing the threshold from 0.7 to 0.9 resulted in more merged groups and more refined groupings, respectively (Table 4.2, Figure 2E).

The reduction ratio analysis indicated that K-Means reduced table comparisons by up to 83%, Hierarchical Clustering by up to 80%, and DBSCAN by up to 77% (Table 4.3, Figure 2F). The variation in reduction ratios across methods is primarily due to parameter configurations rather than inherent limitations of the algorithms, suggesting that all methods are effective in optimizing the process.

**Table 3.** Reduction Ratio by Clustering Method.

| Clustering Method | Reduction Ratio (%) | Figure |
| --- | --- | --- |
| K-Means | Up to 83% | 2F |
| Hierarchical Clustering | Up to 80% | 2F |
| DBSCAN | Up to 77% | 2F |

Overall, the integrated use of clustering and semantic similarity techniques enhanced the efficiency and accuracy of schema matching and entity resolution within the MAUDE database.

### 4.2 Token Efficiency Assessment

A similarity threshold of 0.8 was selected, and the K-Means clustering algorithm was manually configured to specify three clusters. Feature extraction was performed using TF-IDF vectorization, yielding an optimal solution with an F1 score of 1.0. This configuration resulted in the merging of 113 tables into 14 groups. The assessment revealed a significant reduction in token usage due to the merging process. Prior to merging, the prompt text (prompt_before_merging.txt) comprised a total of 113,954 tokens, including 2,167 tokens from context.txt and 111,787 tokens from table descriptions. After merging, the prompt text (prompt_after_merging.txt) contained only 15,831 tokens, with the context remaining constant at 2,167 tokens and the table descriptions reduced to 13,664 tokens. This represents an absolute reduction of 98,123 tokens in table descriptions and a percentage reduction of approximately 87.78%. Consequently, the prompt length now meets the context window requirements of almost all mainstream LLM chatbot interfaces. All prompt texts generated during this process are available for direct use by MAUDE researchers in our open-source GitHub project, enhancing accessibility and facilitating further research.

## 5. Discussion

### 5.1 Effectiveness of Combining Clustering with Semantic Similarity Evaluations

Integrating semantic similarity evaluations via the DeepSeek V2.5 API with clustering algorithms significantly enhanced dimensionality reduction in the MAUDE database. The F1 score improved from 0.51 using clustering alone to 1.00 when combined with semantic assessments, underscoring the critical role of semantic understanding in accurately identifying and merging similar tables. This combined approach not only increased the precision of table merging but also ensured comprehensive recall, effectively minimizing both false positives and false negatives.

### 5.2 Computational Efficiency and Cost Analysis

The initial clustering phase, supplemented by Jaccard similarity-based prefiltering, reduced the number of table pairs requiring semantic similarity computations by 77% to 83%. This reduction is crucial for managing computational resources, especially when dealing with large-scale databases with numerous potential table pairs. Although utilizing the DeepSeek V2.5 API introduces computational overhead, the significant gains in clustering accuracy justify this investment. Achieving near-perfect F1 scores with fewer than 1,425 API calls demonstrates the cost-effectiveness of the proposed methodology, particularly in contexts where manual table merging is impractical due to dataset size or complexity.

### 5.3 Advantages of the Proposed Methodology

By combining structural similarities (via the Jaccard index) with semantic similarities (via the open-source DeepSeek V2.5 API), the methodology offers a comprehensive assessment of table relationships. This dual approach captures both overt structural overlaps and nuanced semantic relationships, facilitating more accurate and meaningful table merging. The success of this methodology in the MAUDE database, despite the absence of standardized classifications, indicates its scalability and adaptability to other domains with complex, unstandardized datasets. Systematic documentation of configurations and methodological steps enhances reproducibility, allowing other researchers to validate and extend the approach.

### 5.4 Impact on Context Window Limitations

Compressing nearly 80% of the table descriptions enabled the consolidation of representative table structures and sample data, as well as the inclusion of official guideline texts, within the context window limits of advanced language models. Specifically, the compressed data fits within a 128k token context window—the maximum for certain APIs—and even within a 32k token limit for chatbots. This compression is pivotal for leveraging Large Language Models (LLMs) in data analysis, as it ensures that the models can process the entire dataset without exceeding token limitations. By maintaining the integrity of the data within these context windows, the methodology enhances the effectiveness of LLMs in generating accurate and comprehensive insights.

### 5.5 Comparison with Manual Grouping

The automated methodology closely aligned with manual groupings provided by domain experts, as evidenced by an F1 score reaching up to 1.00. This alignment validates the effectiveness of the proposed approach in replicating expert judgment, reducing reliance on time-consuming manual processes. The high consistency between the automated and manual groupings underscores the reliability of combining clustering with semantic similarity evaluations.

### 5.6 Limitations and Future Directions

While the DeepSeek V2.5 API is open source and offers robust semantic similarity evaluations, reliance on any specific tool introduces considerations regarding maintenance and updates. Future work could explore integrating additional open-source semantic similarity tools or developing domain-specific models to enhance flexibility. Semantic ambiguities inherent in complex datasets like MAUDE may still pose challenges; incorporating domain-specific ontologies or knowledge graphs could further improve semantic disambiguation and similarity assessments. Expanding the methodology to include additional dimensionality reduction techniques, such as autoencoders^33^ or manifold learning methods^34^, could enhance robustness and scalability. Empirical validation across diverse domains is necessary to substantiate the methodology’s effectiveness and adaptability.

### 5.7 Implications for Medical Informatics and Beyond

The successful application of this automated dimensionality reduction methodology in the MAUDE database has significant implications for medical informatics. By improving data quality and analysis efficiency, it facilitates more reliable insights into patient safety events, supporting regulatory decision-making and enhancing patient outcomes. The ability to compress extensive datasets within the context window limits of LLMs unlocks new potentials for advanced data analysis and interpretation. The methodology’s potential applicability extends beyond healthcare, offering a scalable framework for data management and analysis in industries characterized by complex and unstandardized datasets. Automating the identification and merging of similar tables streamlines data processing workflows and ensures consistency in data representation, enabling researchers and practitioners to focus on higher-level analytical tasks.

## 6. Conclusion

This study presents a novel automated methodology for dimensionality reduction of complex databases, demonstrated through the FDA’s MAUDE database. By integrating clustering algorithms with semantic similarity evaluations using the DeepSeek V2.5 API, we successfully reduced the number of tables from 113 to approximately 14–16 groups. This significant reduction enhances data quality and improves analysis efficiency, making the dataset more manageable for both manual analysis and advanced analytical tools such as Large Language Models (LLMs).

The combination of clustering and semantic similarity evaluations proved highly effective, increasing the F1 score from 0.51 with clustering alone to a perfect score of 1.00. This improvement underscores the importance of semantic understanding in accurately identifying and merging similar tables. Additionally, the methodology achieved computational efficiency by reducing the number of table pair comparisons by up to 83% through initial clustering and prefiltering steps.

Our findings demonstrate that the methodology closely aligns with expert manual groupings, validating its reliability and effectiveness. The significant reductions in required computations make the approach both cost-effective and scalable. Moreover, by compressing the data to fit within the token limitations of LLMs, the methodology preserves contextual integrity, enabling more accurate and comprehensive analyses. While this approach was applied to the MAUDE database, its adaptability to other industries with complex and unstandardized datasets highlights its versatility.

Future research will focus on enhancing the methodology’s adaptability and robustness by integrating additional semantic similarity tools, exploring alternative dimensionality reduction techniques, and applying the approach to diverse datasets across various domains. By streamlining data processing workflows and ensuring consistency in data representation, this methodology holds substantial potential for improving data analysis efficiency and accuracy in multiple fields. In conclusion, the proposed automated dimensionality reduction methodology effectively addresses the challenges associated with large, complex databases, facilitating more efficient data processing and enabling compatibility with advanced analytical tools. This advancement is poised to significantly impact data analysis workflows, regulatory decision-making, and ultimately, patient outcomes in the healthcare industry and beyond.

## Data Availability

All data produced are available online at https://github.com/leiMizzou/Maude-Schema-Analysis

## 8. Conflict of Interest

The authors declare no conflict of interest.

## 9. Funding

This research was supported by Fund Program for the Scientific Activities of Selected Returned Overseas of Professionals in Shanxi Province under Grant No. 20230039. The funding agency had no role in the study design, data collection and analysis, decision to publish, or preparation of the manuscript.

## 10. Data Availability

The data supporting the findings of this study are available from the FDA’s MAUDE database at [https://www.fda.gov/medical-devices/maude-database](https://www.fda.gov/medical-devices/maude-database) upon reasonable request. All other data generated or analyzed during this study are included in this published article and its supplementary information files.

## 11. Supplementary Material**

## Appendix A Grouping of MAUDE and Self-Built Statistical Tables

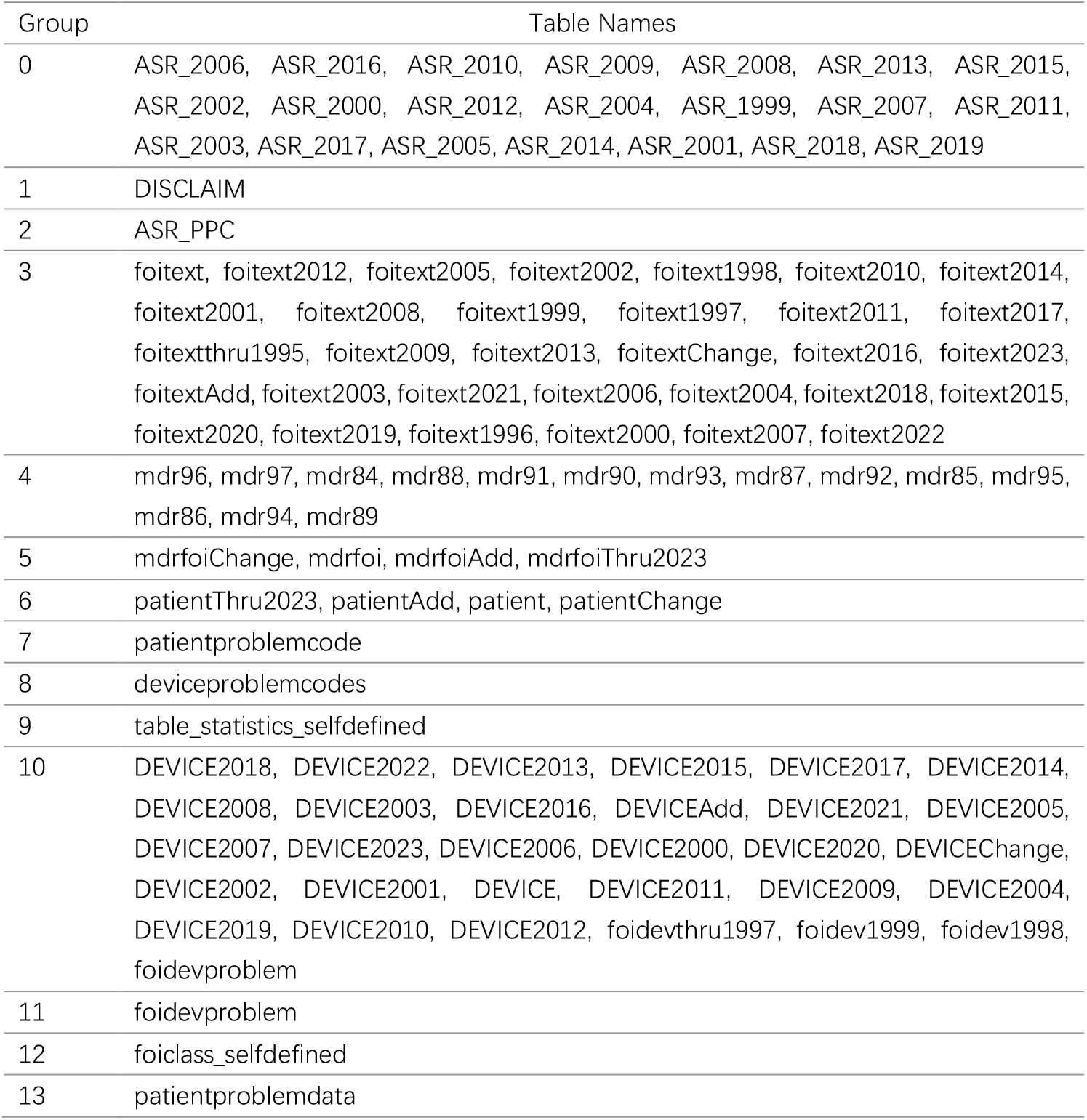

## Footnotes

i Publicly available and can be downloaded from the FDA MAUDE Database.https://www.fda.gov/medical-devices/medical-device-reporting-mdr-how-report-medical-device-problems/mdr-data-files

## Notes

### Competing Interest Statement

The authors have declared no competing interest.

### Funding Statement

Fund Program for the Scientific Activities of Selected Returned Overseas Professionals in Shanxi Province

### Author Declarations

U.S. Food and Drug Administration (FDA). Manufacturer and User Facility Device Experience (MAUDE). 2024. Accessed November 1, 2024. https://www.fda.gov/medical-devices/mandatory-reporting-requirements-manufacturers-importers-and-device-user-facilities/about-manufacturer-and-user-facility-device-experience-maude-database

